# Graph-based stochastic modelling of glioblastoma invasion using patient-specific structural brain connectomes

**DOI:** 10.64898/2026.07.22.26358652

**Authors:** Martin Kukrál, Roy A.M. Haast, Irena Holečková

## Abstract

Glioblastoma (GBM) is the most common and aggressive primary malignant brain tumor in adults with extremely poor prognosis. Complete surgical treatment is practically impossible, as the true extent of GBM infiltration cannot be fully delineated using currently available *in vivo* neuroimaging methods, leading to frequent recurrences and low overall survival. Consequently, mathematical models are being developed to estimate the GBM expanse beyond the visible tumor mass, providing additional information for treatment planning and patient prognosis. Here, a novel graph-based stochastic mathematical model of GBM invasion using patient-specific structural brain connectome data is proposed. The model is assessed using publicly available UCSF-PDGM dataset to demonstrate GBM invasion dynamics across multiple patients and anatomical locations. Additional scaling using fractional anisotropy (FA) is tested and evaluated. Parameter sensitivity analysis is provided to explore model’s behavior under different settings. Ablation testing is performed to suppress model mechanisms utilizing the structural connectome, showing that the tentacle-like extrusions from the tumor core emerge only if the patient-specific connectome is utilized. The model seems to capture GBM micro-infiltration along white matter tracts to a very high degree, making it a potential tool for studying distant recurrences farther from the resection cavity and GBM invasion dynamics in relation to the structural connectome. Full source code is publicly available, ensuring complete transparency of the study.

## 1 Introduction

Glioblastoma (GBM), the most common and aggressive primary malignant brain tumor in adults, remains one of the greatest challenges in modern neuro-oncology due to its extremely poor prognosis despite advances in molecular biology, genomics, and modern neuroimaging techniques (Królikowska *et al*., 2025). The World Health Organization (WHO) classification of central nervous system (CNS) tumors categorizes GBM as the highest grade 4 diffuse glioma subtype, characterized, e.g., by isocitrate dehydrogenase (IDH) wild-type status, molecular and chromosomal alterations, extensive invasion into surrounding brain tissue, microvascular proliferation, necrosis, and highly dynamic tumor microenvironment (TME) (Singh *et al*., 2025).

GBM treatment aims for an initial maximal safe resection, followed by targeted radiotherapy and temozolomide chemotherapy (Wen *et al*., 2025). Nevertheless, even for patients receiving standard treatment, the median overall survival (OS) is 15–20 months and only 8–9 months after recurrence (Fortunato *et al*., 2026). Given these poor outcomes, strategies that safely maximize tumor resection remain an important focus of surgical innovation. Surgery guided by multimodal brain mapping aims for a complete resection of the tumor outlined by contrast-enhanced magnetic resonance (MRI) without damaging eloquent areas, but increasing the extent of resection (EOR) beyond the contrast-enhancing tumor into the noncontrast-enhancing tissue was shown to be beneficial in multiple patient subgroups (Gerritsen *et al*., 2022). However, current neuroimaging techniques used to delineate infiltration margins fail to precisely capture GBM micro-infiltration due to their limited ability to resolve microstructural differences, so even surgeries with increased EORs lead to recurrences (Zhu *et al*., 2024; Dada *et al*., 2026). Consequently, mathematical models of GBM invasion are seen as a complement to imaging data capable of estimating heterogeneous spatial distributions of cancer cell infiltration beyond the visible tumor mass, signaling the need to develop data-driven and patient-specific modelling approaches to guide personalized treatment decisions (Zhang *et al*., 2025).

I would This work proposes a novel graph-based stochastic mathematical model of macroscopic GBM invasion along patient-specific structural brain connectomes, reconstructed from *in vivo* diffusion MRI data. The structural connectome is represented as a graph structure constructed from fiber tract reconstructions obtained from advanced tractography methods. The mathematical model then uses this graph to simulate patient-specific GBM growth.

## 2 Related works

Over the years, many conceptually distinct glioma and specifically GBM modelling approaches have been formulated at both macroscopic and microscopic scales, utilizing various molecular and imaging data obtained via *in vivo* and *in vitro* experiments (Falco *et al*., 2021). The majority of macroscopic models represent GBM invasion as a kind of reaction-diffusion system described using partial differential equations (PDEs). An alternative to reaction-diffusion PDEs is presented by discrete agent-based systems that typically model GBM spread at the cellular level, causing them to incorporate stochasticity to a greater extent than more aggregative continuum models (Jørgensen *et al*., 2023). Given that the proposed model is macroscopic, the focus is placed on the continuum models.

Early reaction-diffusion models were developed in the 1990s, with later work by Swanson *et al*. (2000) providing a widely adopted formulation and refinement of the original model by accounting for differences in the diffusivity of glioma cells in white and gray matter. This idea of heterogeneous diffusion was expanded by Jbabdi *et al*. (2005), originally for low-grade gliomas, who introduced anisotropic spread by replacing diffusion coefficients with diffusion tensors derived from diffusion tensor imaging (DTI), an MRI technique that has many uses in neurosurgery, notably for non-invasive mapping of white matter tracts (Costabile *et al*., 2019).

Around the same time, Clatz *et al*. (2005) have also worked with DTI, while introducing biomechanical components reflecting the effect of tumor mass on the local environment using a linear elastic brain constitutive equation. Bondiau *et al*. (2008) also incorporate the effect of tumor mass on surrounding tissue, but through a mechanical component coupled with a diffusion component, which are complementary and can be tuned independently. Hogea *et al*. (2008) provided a conceptually similar approach, but using a system of coupled nonlinear PDEs. More recently, Lipková *et al*. (2022) proposed a model that incorporates physical pressure directly derived from tumor dynamics and patient-specific anatomy, allowing estimations of critical conditions such as intracranial hypertension and brain midline shift, with the potential to evaluate neurological and cognitive impairments.

Other models focus on the molecular heterogeneity of GBM. Wise *et al*. (2008) proposed a two-population diffuse interface model that describes the dynamics of dead and viable tumor cells. A three-population model provided by Martínez-González *et al*. (2012) consists of normoxic tumor cells, hypoxic tumor cells, and necrotic tissue, whose interaction leads to the emergence of pseudopalisades, which are described as hypercellular regions surrounding the central tumor necrosis. Gerlee and Nelander (2012) developed a stochastic model that incorporates phenotypic switching between proliferative and migratory states, which is formulated for multidimensional lattices and also as a continuum model. Cai *et al*. (2016) propose a coupled system that considers four tumor cell phenotypes, which is aimed at investigating GBM growth in response to dynamic changes in the chemical and hemodynamic microenvironment. Patel and Hathout (2017) also focus on the effect of necrotic tissue by formulating a necrosis term that is activated once cell density has surpassed tissue support capacity. Recently, Tursynkozha *et al*. (2025) described a model that captures the three-layer structure of multicellular spheroids during avascular tumor growth using subpopulations of migratory, proliferative, and necrotic cells.

The go-or-grow hypothesis, which states that highly migratory cells have a lower proliferation rate compared to actively proliferating cells that move slowly (Alfonso *et al*., 2017), has significantly influenced the mathematical modeling of GBM. Many of the models mentioned above use it as an argument for the formulation of multiple cell subpopulations. Thiessen *et al*. (2025) provide an overview of this class of models, as well as their generalized formulation.

Future directions towards increased stochasticity, the inclusion of more advanced neuroimaging methods, and machine learning (ML) were suggested by Cozzi *et al*. (2025). Although ML algorithms can deliver exceptional results in various clinical tasks (Tbahriti *et al*., 2025), in GBM invasion modelling, their role remains mostly supportive, the main applications being parameter estimation or hybrid modelling schemes. A brief overview of various ML-based extensions of mathematical models is provided by Kukrál (2026).

Although tractography was previously used to estimate the extent of GBM invasion by Kis *et al*. (2022) and Abbasian *et al*. (2025), to our knowledge, it was not previously used directly in a mathematical model. Therefore, the proposed graph-based stochastic model is considered a novel methodological approach that explicitly describes the GBM invasion pathways and dynamics in relation to the structural brain connectome. The framework models GBM micro-infiltration along white matter tracts to a degree unseen in previous models, making it a potential tool for surgery and radiotherapy planning, studying GBM recurrences located farther from the resection cavity, and for exploring the interactions between GBM and structural brain connectome.

## 3 Materials and methods

The GBM invasion modelling consists of three phases: the initial processing of patient-specific diffusion MRI data, the reconstruction of fiber tracts comprising the structural brain connectome, and the iterative computation of the graph-based stochastic mathematical model using the newly created graph structure to estimate GBM spread in subsequent time steps. The connectome reconstruction and mathematical model are fully independent, making the proposed approach modular and flexible on the basis of available data and its properties. Every step of the pipeline is implemented in Python and made available in the associated GitHub repository, providing a framework for additional experiments. The methodology is shown in Figure 1.

**Figure 1.**
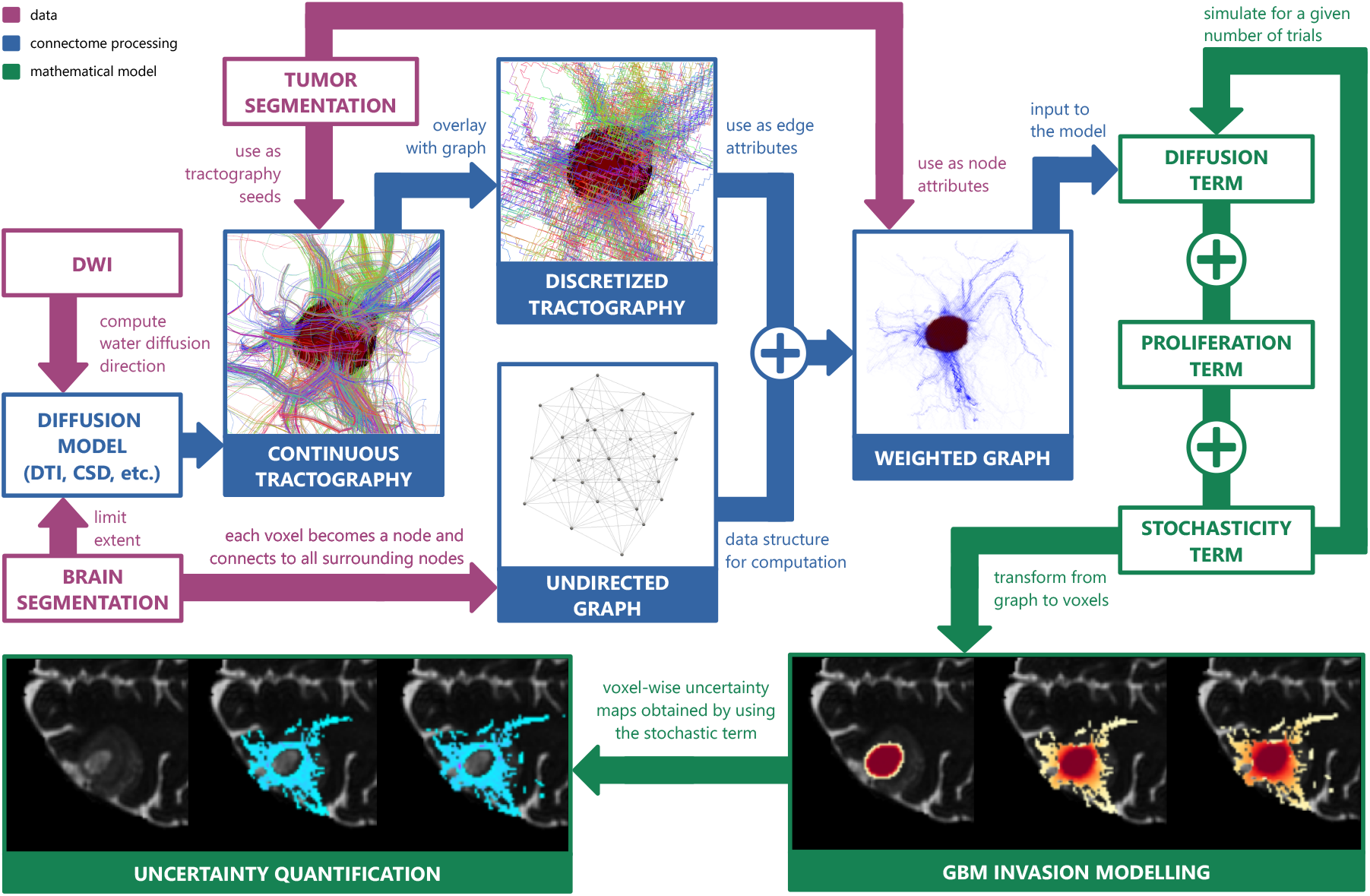
Illustration of the proposed GBM invasion modelling methodology. Abbreviations: CSD (constrained spherical deconvolution), DTI (diffusion tensor imaging), DWI (diffusion-weighted imaging), GBM (glioblastoma).

### 3.1 Data

To demonstrate the proposed mathematical model, simulations are performed using the publicly available University of California San Francisco Preoperative Diffuse Glioma MRI (UCSF-PDGM) dataset by Calabrese *et al*. (2022a,b), which can be freely downloaded from The Cancer Imaging Archive (TCIA) through IBM Aspera Connect. The dataset contains a total of 495 unique subjects with histopathologically confirmed adult-type diffuse gliomas, mostly GBMs, obtained between 2015 and 2021 using a 3T scanner, a dedicated 8-channel head coil, and a single preoperative MRI imaging protocol. UCSF-PDGM provides data from various MRI modalities and multiple segmentation masks in the Neuroimaging Informatics Technology Initiative (NIfTI) format.

Diffusion-weighted imaging (DWI) data contains a b0 image and 55 diffusion gradients sampled under a fixed *b*-value (*b* = 2, 000 s*/*mm^2^), making this a single-shell high-angular-resolution diffusion imaging (HARDI) data. The authors of the dataset state that the DWI data have been Eddy current corrected with outlier replacement. Tumor segmentations and brain masks are provided.

The MRI data in the UCSF-PDGM dataset, excluding DWI, is resampled to 1 mm^3^ voxels and spatially aligned using Advanced Normalization Tools (ANTs, available for Python via the wrapper library `antspyx` ), specifically through automated non-linear registration. For the purposes of this study, the registration of DWI to the rest of MRI data was performed using the “`ElasticSyN`” variant, which performs symmetric normalization (SyN) proposed by Avants *et al*. (2008) with mutual information (MI) as a similarity metric, while also introducing elastic regularization (ANTs Contributors, 2017). Resampling interpolation was performed using B-splines.

Taking into account the similarity in contrast between the b0 images and the *T*_2_-weighted images (T2WIs), the optimal transformation was computed using them and applied to all 55 gradient directions, ensuring complete 4D DWI registration. The registered DWI was then skull-stripped using the brain mask. The chosen registration method is similar to the method originally used in the UCSF-PDGM dataset and should be sufficiently flexible, while providing certain deformation constraints.

### 3.2 Connectome processing

To reconstruct the structural brain connectome, the registered DWI data is first used to fit a model expressing the diffusion strength and direction of water molecules, which enables the estimation of fiber tract arrangement due to the well-known fact that water diffuses easier along axons rather than perpendicular to them. DTI is the most commonly used diffusion model for this task, but more advanced alternatives are available, such as constrained spherical deconvolution (CSD) by Tournier *et al*. (2007), which addresses the inability of DTI to express crossing fibers, since 70–90% of human white matter is composed of at least two fiber populations (Dell’Acqua and Tournier, 2019).

In this study, the CSD implementation provided in the Python library dipy by Garyfallidis *et al*. (2014) is used to model water diffusion in brain tissue. The response function is automatically estimated from regions with fractional anisotropy (FA) > 0.7 and the maximal spherical harmonics order is set to *ℓ*_*max*_ = 8. The resulting fiber orientation distribution functions (fODFs) are then used by a deterministic tractography algorithm to estimate the fiber tracts surrounding the tumor mass, which was obtained using tumor segmentation labels as *necrotic tumor* ⋃ *enhancing tumor*. The GBM tumor mass was also used to generate initial seeds for tracking with a density of 1 seed per tumor voxel, which means that the reconstructed subset of the brain connectome corresponds to tracts that are directly connected to the GBM tumor mass. Additional parameters were set as follows: tracking step size at 0.2 mm, maximum angle at 20^°^, minimum streamline length at 2 points, maximum streamline length at 1,000 points, and maximum number of iterations for deconvolution convergence at 50. Reaching voxels with generalized fractional anisotropy (GFA) < 0.25 was used as a stopping criterion. In this way, relatively weak local connections around the GBM mass are preserved without disrupting dense and far-reaching tracts.

During tractography, the estimated structural connectomes were situated in real-world coordinates using an associated affine matrix **A**. Consequently, the 3D points comprising the streamlines can be re-mapped back to the voxel space by the inverse affine matrix **A**^−1^. Once the transformation is complete, the tract discretization procedure begins by rounding the coordinates to the nearest integer values, making them correspond to indices in the voxel grid. Because this operation can disrupt the continuity of the original streamlines, it is necessary to clean their discrete counterparts. Firstly, identical consecutive points are replaced by just one point. Secondly, if the consecutive points are not adjacent in the full 3D neighborhood (26-connectivity), they are reconnected through the shortest path obtained using the Chebyshev distance by iteratively adapting the coordinates of the first point one step at a time until it converges with the second point. Discrete tracts comprised of only one point are discarded.

Finally, the graph can be assembled. The overall graph structure is created using the binary brain mask by initializing graph nodes in the center of voxels that correspond to brain tissue and connect all neighborhoods (26-connectivity) via undirected edges. Node weights are determined using the overlapping tumor mask segmentation (1 ∼ tumor, 0 ∼ no tumor), which can be first smoothed to remove the non-physiological hard edge at the tumor boundary. Edge weights are computed by counting all discretized tract streamlines that pass through a given pair of nodes. It is assumed that the original voxel dimensions are isotropic.

Because the tractography was initialized from 1 seed per tumor voxel, no additional normalization is required. Otherwise, seed density must be taken into account by decreasing the edge weight proportionally to the number of tracking seeds. In `dipy`, the seed parameter is treated as the number of seeds to place along each dimension (DIPY developers, 2026). Given that the graphs are defined in a 3D space, the following normalization is performed:

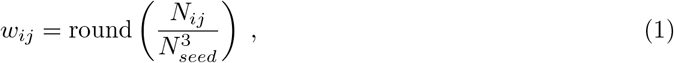

where *w*_*ij*_ is the edge weight between nodes *v*_*i*_, *v*_*j*_, corresponding to the seed-normalized number of passing streamlines rounded to the nearest integer, *N*_*ij*_ is the unnormalized number of passing streamlines, and *N*_*seed*_ is the value of the seed parameter given to the `dipy` function. If *N*_*seed*_ = 1, then *w*_*ij*_ = *N*_*ij*_.

Edge weights can be further adjusted, for example, on the basis of tract integrity. Here, we test relative scaling using FA, which is well-known and commonly used as a quantitative white matter integrity metric. The goal is to utilize the difference between local FA and the overall median FA, essentially amplifying or reducing the edge weights to model the relative preferential spread of GBM tumor cells along the tracts with perceived higher integrity. Although Figley *et al*. (2022) correctly points out many limitations of FA as a biomarker, it can still be suitable for scaling, as the goal is to relatively scale the edges rather than to interpret FA values in absolute terms. Specifically, the relative scaling is computed as follows:

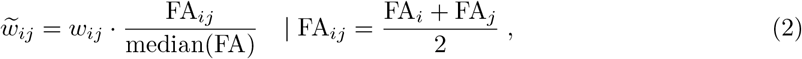

where 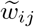 is the scaled edge weight, FA_*i*_ is the FA value at node *v*_*i*_, FA_*j*_ is the FA value at node *v*_*j*_, and FA_*ij*_ is their average assigned to the edge that connects them. Because the graph overlaps with the image space, the FA values at each node can be obtained from the corresponding voxels. Naturally, other metrics could be used instead of FA, as well as other scaling approaches.

The assembled weighted graph serves as a model that can be interpreted biologically. The node weights represent the fraction of tissue occupied by the GBM tumor in a given voxel. Edge weights express the degree of axonal connectivity between two adjacent voxels, which is crucial for GBM invasion, as GBM has been quantitatively shown to preferentially spread along white matter tracts, which seem to guide tumor cell migration as a sort of latent scaffolding (Shimizu *et al*., 2026). Using undirected edge weights assumes that the GBM motility is equal in both directions.

The mathematical model is fully independent of brain connectome processing. Therefore, it is possible to use other tractography methodologies, as long as they produce 3D reconstructions of fiber tracts that can be used to add edge weights to the graph. Similarly, other tumor segmentation approaches can be used, such as initializing different node weights to necrotic and enhancing tumor tissue. Furthermore, the pipeline can be limited only to the brain tissue closest to the GBM tumor mass to conserve computational time and memory usage. The graph construction is summarized in Figure 2.

**Figure 2.**
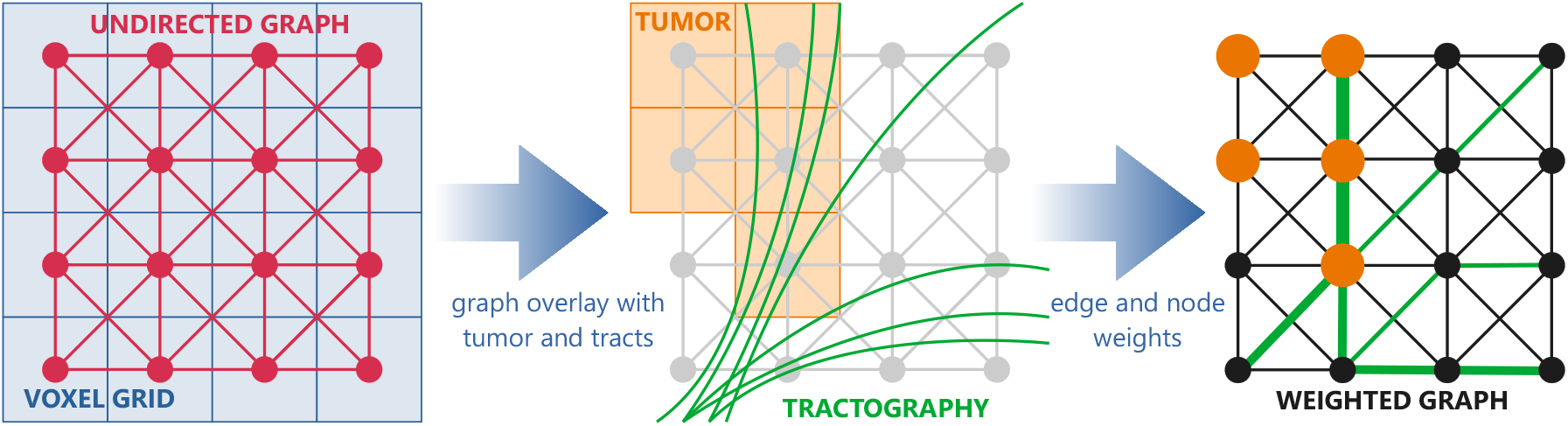
Simplified illustration of 2D weighted graph construction using tumor segmentation and tractography. In reality, the procedure is applied to 3D data.

### 3.3 Mathematical model

The proposed mathematical model of GBM invasion is conceptually derived from the original reaction-diffusion PDE provided by Swanson *et al*. (2000) as:

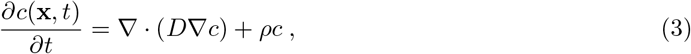

where *c* = *c*(**x**, *t*) represents the number of tumor cells in position **x** ∈ ℝ^3^ and at time *t, D* [cm^2^*/*day] is the diffusion coefficient of tumor cells, *ρ* [1*/*day] is the proliferation rate (net growth rate of tumor cells including proliferation and death), ▽ is the gradient operator, and ▽· is the divergence operator. The first term describes the motility of tumor cells and the second term describes their growth.

Let *G* = (*V, E*) be a graph obtained from connectome processing, which is composed of a set of nodes *V* = { *v*_1_, *v*_2_, …, *v*_*n*_ } for *n* = |*V*| and a set of undirected edges *E* ⊆ {{*v*_*i*_, *v*_*j*_} : *v*_*i*_, *v*_*j*_ ∈ *V, i* ≠ *j* }, weighted by a function *ω* : *E* → ℕ, assigning the number of passing tractography streamlines (1), and optionally also by 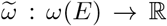, which additionally scales the edge weights (2). Then, the graph-based GBM invasion stochastic differential equation (SDE) can be initially outlined as:

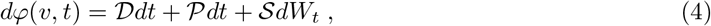

where *φ* = *φ* (*v, t*) ∈ [0, 1] is the fraction of tissue occupied by the GBM tumor at a graph node *v* ∈ *V* (i.e., the node weight) and time *t, W*_*t*_ is the value of a Wiener process at time *t, D* is the diffusion term, *P* is the proliferation term, and *S* is the stochasticity term.

The diffusion term *D* reflects the motility of tumor cells in *G*, similarly to the first term of the original reaction-diffusion PDE (3). However, instead of providing diffusion strength directly using a diffusion coefficient, it is implicitly contained in the edge weights, with 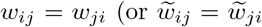 if additional scaling is used) due to the fact that the edges are undirected. The Laplace operator is a second order differential operator interpretable as a divergence of a gradient (note the parallels with equation (3)), with a discrete counterpart that can be used for graphs (Zámečníková and Perfilieva, 2022):

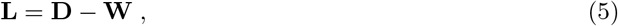

where **L** ∈ ℝ ^*n*×*n*^ is the Laplacian matrix computed using a degree matrix **D** ∈ ℝ ^*n*×*n*^ and a weight matrix **W** ∈ ℝ ^*n*×*n*^, both given as:

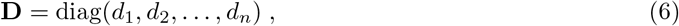

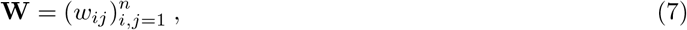

where *d*_*i*_ = ∑ _*j*_ *w*_*ij*_ is the node degree of node *v*_*i*_ (modified from Xu *et al*. (2020)). **L** is deliberately kept unnormalized, as both normalized and random walk Laplacians rescale the weight matrix by the degree matrix (**D**^−1*/*2^**WD**^−1*/*2^ or **D**^−1^**W**) (Xu *et al*., 2020), making the connectivity node-degree relative instead of reflecting absolute tractography streamline counts. Nodes with large node degrees are anatomically meaningful as hubs with strong structural connectivity, which are important for GBM spread, and so removing them would distort the underlying anatomy.

Reaction-diffusion models can directly encode the go-or-grow dichotomy at the macroscopic level, typically by featuring non-linear diffusion as a response to density-dependent rates, various environmental factors, or the crowding effect, which reduces the migratory capacity of GBM cells (Thiessen *et al*., 2025). Given the graph-based formulation of the proposed model, the effect of tumor density is integrated by scaling **L** proportionally to *φ* using a decay function *ζ* (*φ* ; Θ _*ζ*_) : [0, 1] → [0, 1] with a set of parameters Θ _*ζ*_. Here, two specific decay functions are investigated:

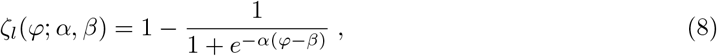

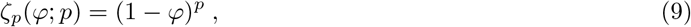

where *ζ* _*l*_ is a logistic-style decay function with steepness *α* ∈ ℝ_>0_ and inflection point in *β* ∈ (0, 1), and *ζ* _*p*_ is a power-style decay function with exponent *p* ∈ ℝ _>1_. The functions are visualized in Figure 3a. The main difference between both decay functions is the preservation of full diffusion for low-*φ* regions, which is meant to amplify the go-or-grow dichotomy by making the invasive front spread significantly faster than the saturated core regions. The decay matrix **Z** ∈ [0, 1]^*n*×*n*^ is assembled as:

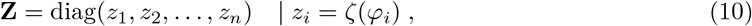

where *φ*_1_, *φ*_2_, …, *φ*_*n*_ are the fractions of tissue occupied by the GBM tumor on all *n* graph nodes. The density-scaled Laplacian 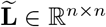 is then given as:

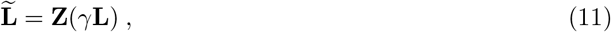

where *γ* ∈ (0, 1] the diffusion scaling factor, conceptually corresponding to a degree of structural connectome utilization by the tumor. The scaling by **Z** is performed row-wise, introducing *φ*-dependent non-linear diffusion. Importantly, 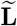 is no longer symmetric. Finally, the diffusion term *D* is assigned as (notation modified from Estrada (2024)):

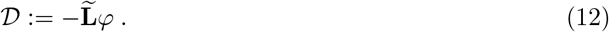

**Figure 3.**
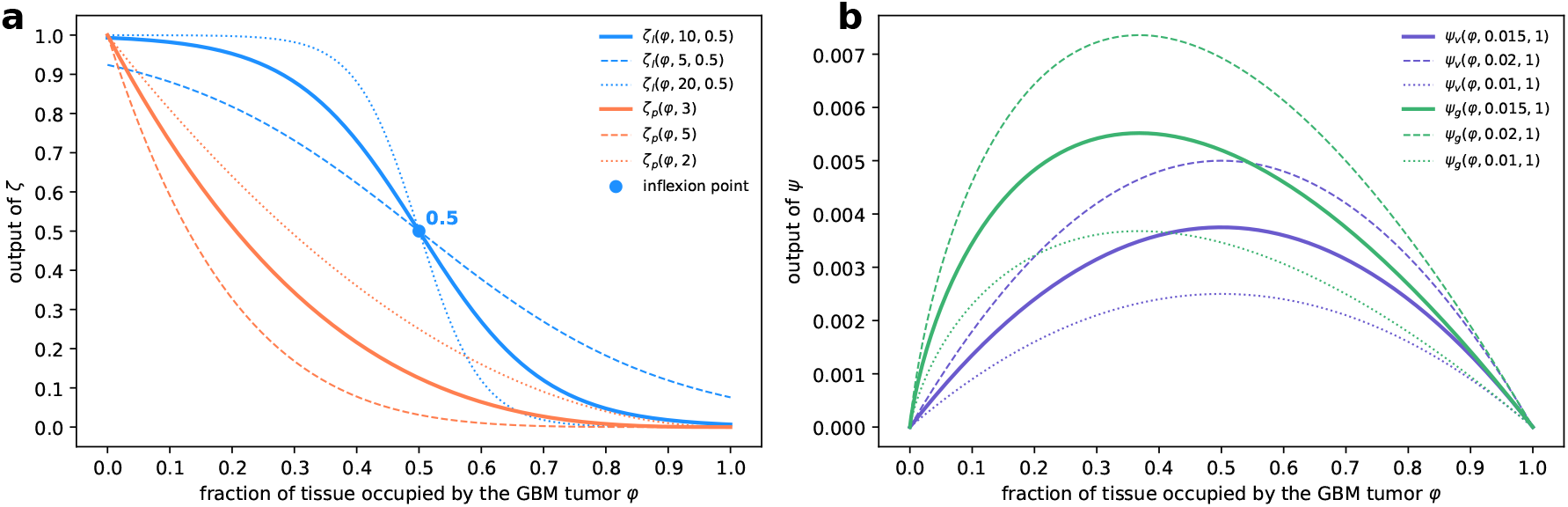
Investigated decay (**a**) and growth (**b**) functions with illustrative parameters.

The diffusion term can be disassembled to show why the Laplacian is density-scaled by **Z** from the left and not from the right. Assuming *γ* = 1, setting all edge weights to 1, and omitting the minus sign for simplicity, following the associative property of matrix multiplication, it can be shown that:

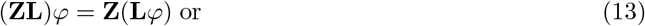

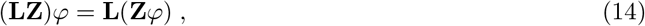

so, the Laplace operator of *φ* at node *v*_*i*_ is (modified from Zámečníková and Perfilieva (2022)):

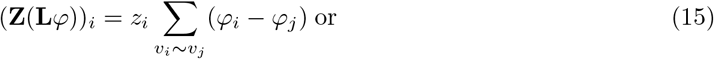

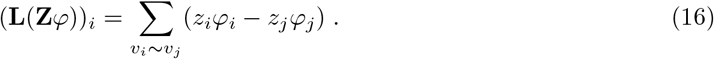

By multiplying from the left, the local diffusion at each node is scaled, whereas multiplying from the right scales the transported mass. Consequently, the utilized transport is no longer pure conservative mathematical diffusion, but rather non-conservative diffusion. Non-conservative diffusion conceptually entails additional interactions with the environment, such as fluctuations of the transported mass during transport (Estrada, 2024). This is very important for GBM modelling, as some tumor cells are expected to naturally die when migrating due to the immune system response, but also proliferate, as the go-or-grow behavior is not strictly binary and tumor cell division has also been shown to occur during migration (Ratliff *et al*., 2023).

If the decay is linear or lies above the corresponding linear decay for larger *φ* values, the invasion pattern is retained, but the tumor core diffuses away and dilutes in the surrounding tissue, which is not physiologically plausible. For this reason, the density scaling is not optional and cannot be arbitrary. This fact may seem to imply that GBM must diffuse non-linearly and dependently on local tumor concentrations, but the observed behavior can simply be a consequence of the model’s numerical properties and not necessarily the GBM pathophysiology.

The proliferation term *P* is conceptually identical to the second term in the original reaction-diffusion PDE (3). Let *ψ* (*φ*; *ρ, κ*) : [0, 1] → [0, 1] be a growth function that governs the proliferation of the GBM tumor, while *ρ* ∈ ℝ is the proliferation rate and *κ* ∈ ℝ_>0_ is the capacity of the environment, i.e., a voxel. In this study, two commonly used growth functions are investigated:

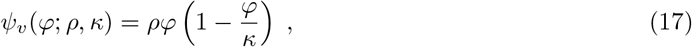

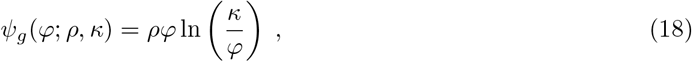

where *ψ*_*v*_ is the Verhulst (or logistic) law and *ψ*_*g*_ is the Gompertz law. The functions are visualized in Figure 3b. Exponential growth is not included, as *φ* is a fraction that cannot grow beyond 1. After omitting *κ* = 1 for simplicity, the proliferation term *P* is assigned as:

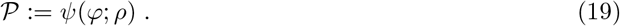

The stochasticity term *S* introduces uncertainty into the model. The previously mentioned Weiner process *W*_*t*_ has time increments with specific properties, such as stationarity, independence, and Gaussian distribution, i.e., *W*_*t*+Δ*t*_ − *W*_*t*_ ∼ *N* (0, Δ*t*) for time *t* and time step Δ*t* (Florescu, 2015). In order to capture random fluctuations in GBM growth due to complex or unknown environmental factors, the environmental noise imposed on proliferation *P* is assigned to the stochasticity term *S* as (*dW*_*t*_ omitted for consistency with equation (4)):

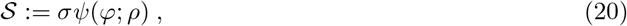

where *σ* ∈ ℝ _≥0_ modulates the intensity of environmental noise (modified notation from Pichór and Rudnicki (2025)). The equation represents multiplicative noise (Volpe and Wehr, 2016). Similar stochastic dynamics were previously explored by Ma *et al*. (2020). Setting *σ* = 0 deactivates the growth stochasticity of GBM and the model becomes deterministic.

By inserting the diffusion (12), proliferation (19), and stochasticity (20) terms into the initial outline (4), the proposed SDE is given using the above defined notation as:

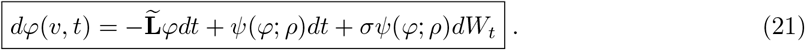

Running the model *k* times with a randomly varying stochasticity term yields *k* potential GBM invasion simulations, which can be aggregated to produce voxel-wise distributions of *φ*. These distributions are then used to spatially compute various statistical characteristics, such as means, medians, standard deviations, etc.

### 3.4 Numerical discretization scheme

The SDE is numerically solved using a Lie–Trotter (first-order) operator splitting scheme, which is used to subdivide a complex equation into simpler parts that are solved sequentially (Blanes *et al*., 2024). The sequence of term computation is *D* → *P* → *S*.

The diffusion term step is computed by the backward Euler method approximating:

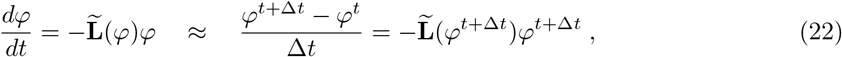

where the dependence of 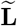 on *φ* is explicitly indicated by adapting a function notation. Rearranging the approximation yields the following:

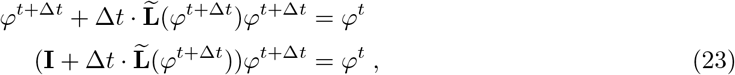

where **I** ∈ { 0, 1 } ^*n*×*n*^ is the identity matrix. To remove high computational costs associated with the iterative solution of a non-linear system, a single Picard iteration is performed, which linearizes non-linear terms by using the solution from the previous time step (Langtangen and Linge, 2017). Specifically, after approximating 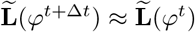, the system matrix **S** ∈ ℝ ^*n*×*n*^ is assigned as:

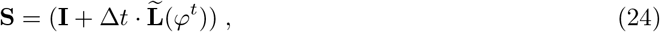

which makes it more straightforward to finally denote the numerical formulation of the future time step as:

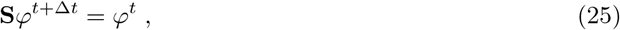

which is solved for *φ*^*t*+Δ*t*^ using the biconjugate gradient stabilized (Bi-CGSTAB) method by van der Vorst (1992) implemented in the scipy library by Virtanen *et al*. (2020), which is designed to numerically solve non-symmetric linear systems. Bi-CGSTAB is a suitable solver, because **S** is non-symmetric as a consequence of 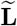 being non-symmetric.

Following the operator splitting scheme and assigning the diffusion step result as *φ*^∗^, the proliferation term step is computed using the forward Euler method as:

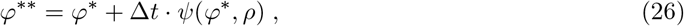

which means that in this implementation, numerically, the GBM tumor cells first diffuse and then proliferate. The multiplicative noise is then incorporated via the Euler–Maruyama Wiener process discretization, the final solution for the next time step is (modified from Higham (2001)):

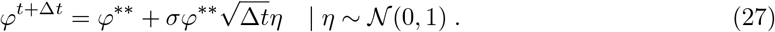

To ensure that *φ* does not impactfully clip out of bounds due to potential floating-point inaccuracies, the values are clipped at the end of each step to the range [0, 1].

## 4 Results

### 4.1 Simulations of glioblastoma invasion

The primary use of the proposed model is to provide patient-specific simulations of GBM invasion dynamics within the brain. To demonstrate the ability of the model to generalize across various cases, four exemplar patients (no. 5, 20, 68, and 92) from the UCSF-PDGM dataset were selected based on three criteria and used to perform the simulations (Figure 4). First, patients had to have wild-type IDH GBM as the final pathological diagnosis in the associated metadata. Second, the GBM tumor mass had to be reasonably sized so that the total volume used for computation could fit into the random-access memory (RAM) of the used computer (Lenovo LOQ 15IRH8 laptop, 16 GB RAM). Third, the GBM tumor masses had to be located in anatomically distinct regions of the brain to show the model’s ability to utilize various subsets of the structural connectome. Given the stochastic nature of the model, the results are shown as voxel-wise medians and interquartile ranges (IQRs).

**Figure 4.**
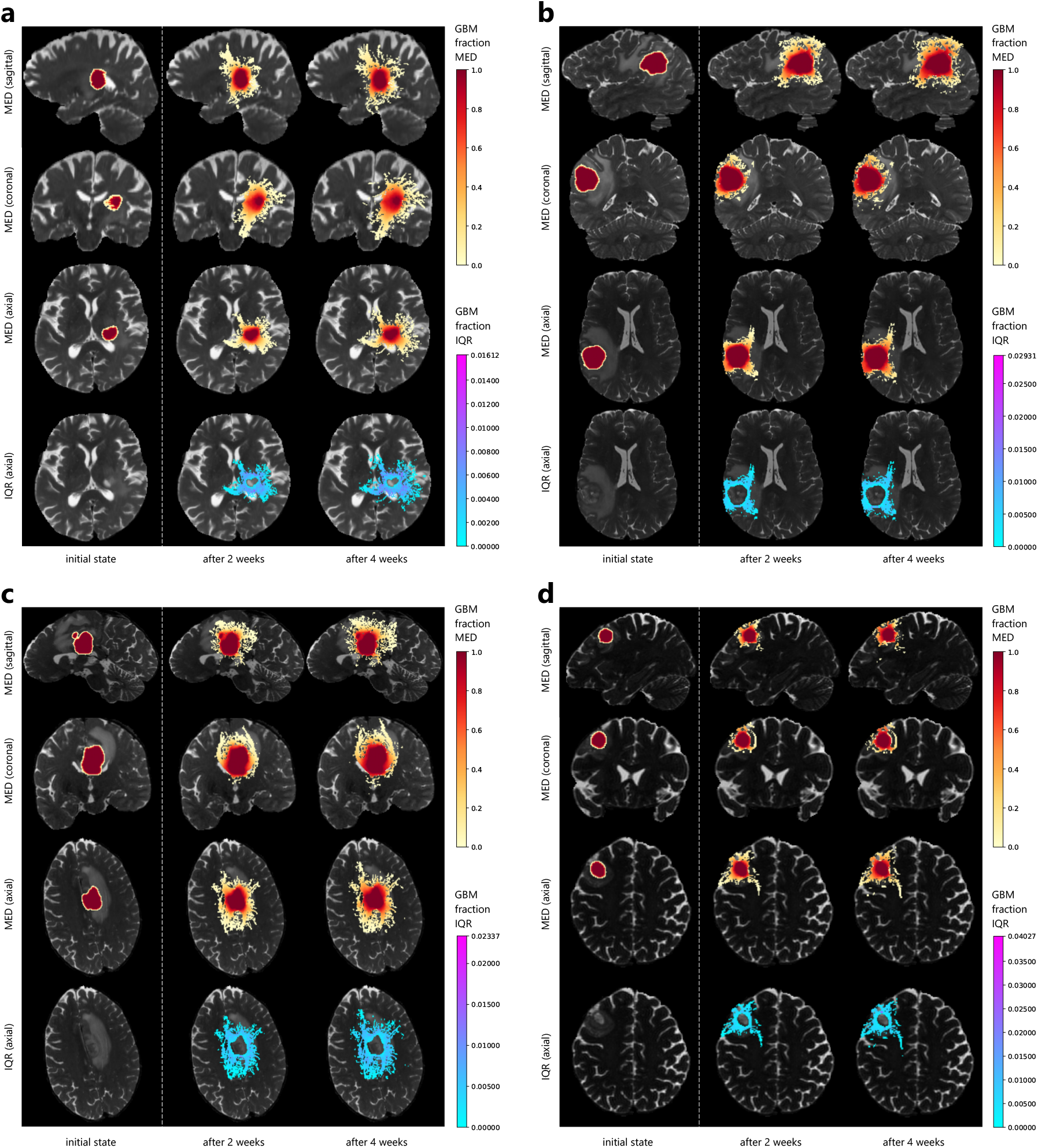
Simulation results overlaid on top of T2WIs: patients no. 5 (**a**), 20 (**b**), 68 (**c**), and 92 (**d**). Coronal and axial slices are scaled to fill more space. No additional FA scaling. Abbreviations: FA (fractional anisotropy), GBM (glioblastoma), IQR (interquartile range), MED (median), T2WI (*T*_2_-weighted image).

The initial GBM tumor masses are located in the following regions: posterior limb of the right internal capsule (patient no. 5), subcortical white matter of the left parietal lobe (patient no. 20), midline involving corpus callosum with superior extension (patient no. 68) and subcortical white matter of the left prefrontal lobe (patient no. 92). Selected examples are located in structurally heterogeneous regions with proximity to a broad range of white matter tracts to explore various invasion patterns.

Patient no. 5 (Figure 4a) shows a preferential GBM spread in both cranial and caudal directions along the corticospinal tract (CST), with an additional major invasion pathway to the left hemisphere through the posterior limb of internal capsule. Further spread is seen laterally along the corona radiata. Patient no. 20 (Figure 4b) exhibits a strong preferential spread along the superior longitudinal fasciculus (SLF), arcuate fasciculus (AF), and possibly other large posterior association fibers. The spread is also modeled above the main tumor mass. Patient no. 68 (Figure 4c) shows tumor infiltration into both CSTs in the cranial direction and also along the central body of the corpus callosum. Patient no. 92 (Figure 4d) shows a more local spread along the U fibers, with a long extension medially along SLF towards deeper white matter structures.

Generally, it can be stated that the model simulates a strong anisotropic invasion of GBM into the surrounding tissue that respects the patient-specific white matter structures. Although not explicitly encoded into the model, two different components of the invading GBM seem to emerge: the invasive front (white-to-yellow) and the tumor core (red-to-orange). The invasive front is far-reaching and might closely reflect the micro-infiltration of GBM cells that would be unobservable via any MRI modality, as the voxel-wise concentrations are still quite low. The degree of uncertainty is comparable between subjects and the maximum IQR was reached by patient no. 92.

The simulation parameters used to obtain the results in Figure 4 were: *γ* = 0.5, the power-style decay function (9) with *p* = 2, the Gompertz growth function (18) with *ρ* = 0.012, and *σ* = 1. The Gompertz growth function was chosen over the Verhulst growth function, as it was shown to better represent empirical tumor growth data (Vaghi *et al*., 2020). The proliferation rate *ρ* was taken from Swanson *et al*. (2000) and other parameters were experimentally chosen to provide interpretable visualizations that seemed realistically plausible. No additional FA scaling was used when processing the structural connectomes. The simulation was performed for 28 days into the future with one state computed per day. The computation extent was limited to a cuboid-shaped region extending 3 cm in all six main directions from the tumor boundary. The tumor edges were smoothed by a Gaussian filter (*σ*_*G*_ = 0.5, kernel size 5 × 5 × 5) to remove the non-physiological hard boundary. Uncertainty was determined by repeating the 28-day simulation 10 times per patient (*k* = 10).

### 4.2 Fractional anisotropy scaling effect

The simulations were repeated with the inclusion of the additional edge weight scaling by FA (2). To inspect the effect of this operation, the difference between scaled and unscaled simulations is provided in Figure 5. All of the parameters used were the same as described previously.

**Figure 5.**
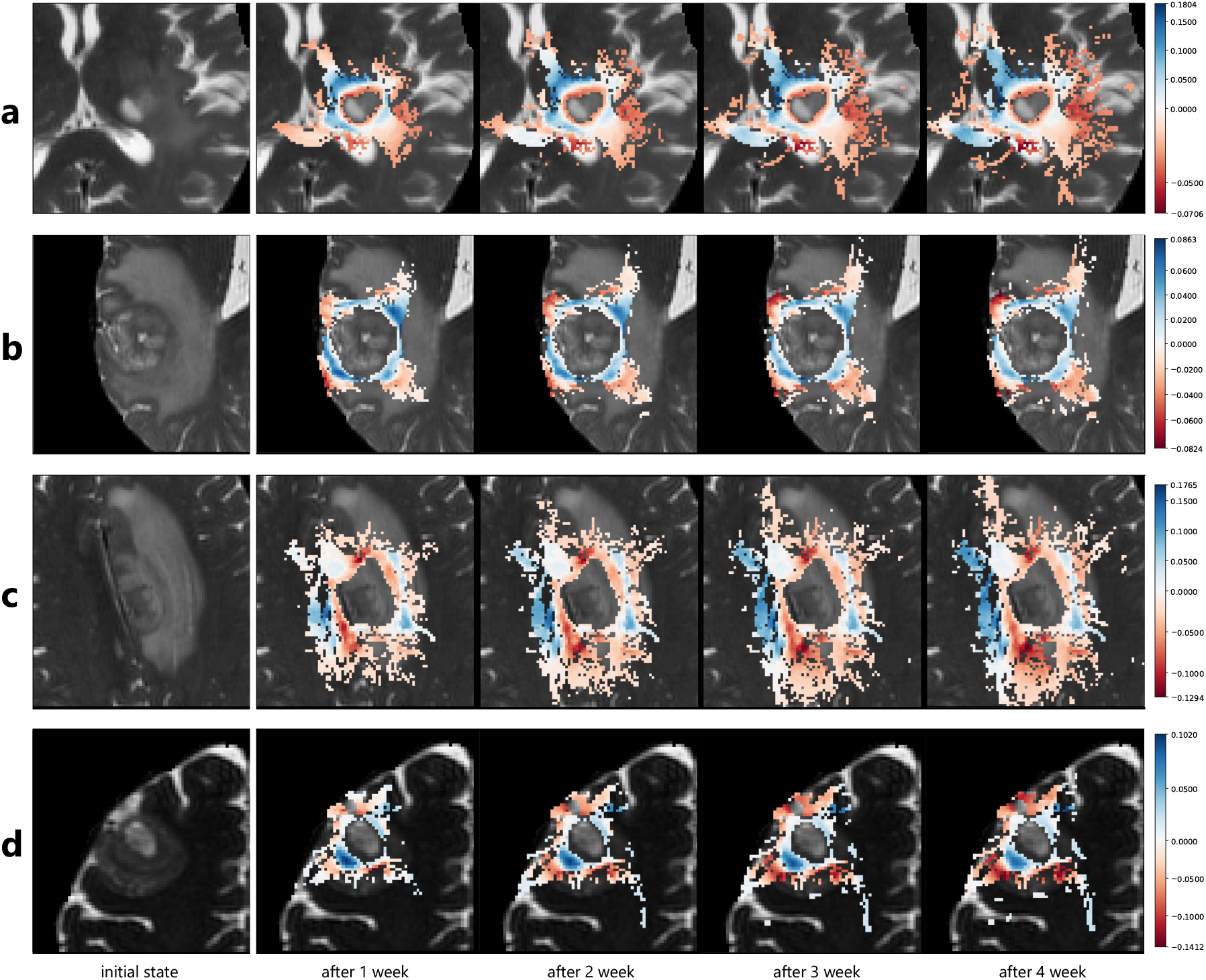
Difference between scaled and unscaled simulations (FA used for scaling) overlaid on top of T2WIs: patients no. 5 (**a**), 20 (**b**), 68 (**c**), and 92 (**d**). Axial slices. Blue indicates region with higher GBM fraction in FA-scaled simulations, red indicates regions with higher GBM fraction in unscaled simulations. Abbreviations: FA (fractional anisotropy), GBM (glioblastoma), T2WI (*T*_2_-weighted image).

After applying the scaling by FA, patient no. 5 (Figure 5a) shows a stronger preference to spread medially into the left hemisphere, while the lateral pathways are slightly suppressed. Patient no. 20 (Figure 5b) does not show a high total difference between both simulation versions, but a slight preference for growth around the tumor mass can be observed when scaling with FA. Patient no. 68 (Figure 5c) shows an increased spread towards the left hemisphere, but also a notable decrease anterior and posterior to the tumor mass. Patient no. 92 (Figure 5d) shows an interesting behavior due to the position of the tumor, as the FA scaling seems to weakly suppress the spread along the U fibers.

### 4.3 Parameter sensitivity analysis

In order to evaluate the effect of various parameters on the GBM invasion dynamics, a sensitivity analysis is performed. In it, one parameter is varied at a time, while the other parameters remain fixed at their default value. The list of analyzed parameters and their tested settings is provided in Table 1. The proliferation rates *ρ* are tested for both Gompertz (18) and Verhulst (17) growth functions (default is Gompertz). When *p* values are tested, the power-style decay function (9) is used, when *α* values are tested, the logistic-style decay function (8) is used with *β* = 0.5 (default is power-style). During analysis, the stochasticity of the model was disabled (*σ* = 0) to obtain deterministic results for parameters *γ, ρ, p*, and *α*.

**Table 1.** Parameters tested in the sensitivity analysis.

| Parameter | Default | Tested parameter values |  |  |  |  |
| --- | --- | --- | --- | --- | --- | --- |
|  |  | 1 | 2 | 3 | 4 | 5 |
| $\gamma$ | 0.5 | 0.05 | 0.25 | 0.5 | 0.75 | 1 |
| $\rho$ | 0.012 | 0.002 | 0.01 | 0.02 | 0.03 | — |
| $p$ | 2 | 2 | 3 | 4 | 5 | — |
| $\alpha$ | — | 5 | 10 | 20 | 30 | — |
| $\sigma$ | — | 0.1 | 1 | 3 | — | — |

To quantitatively evaluate the GBM invasion dynamics, the tumor volume is computed as the sum of all voxels (or, equivalently, nodes). Volume is normalized by the corresponding initial states to yield comparable results in the four exemplar patients. The stochasticity was evaluated using the inter-trial standard deviation (SD) of volumes. During parameter testing, the simulation was computed for 14 days. No additional scaling using FA was performed. Stochasticity was activated when evaluating the effect of *σ*, as it is the parameter that directly affects the degree of randomness, with the number of trials set to 5. The results of the sensitivity analysis for deterministic parameters are provided in Fig. 6a-c and in Fig. 6d for the stochastic parameter *σ*.

**Figure 6.**
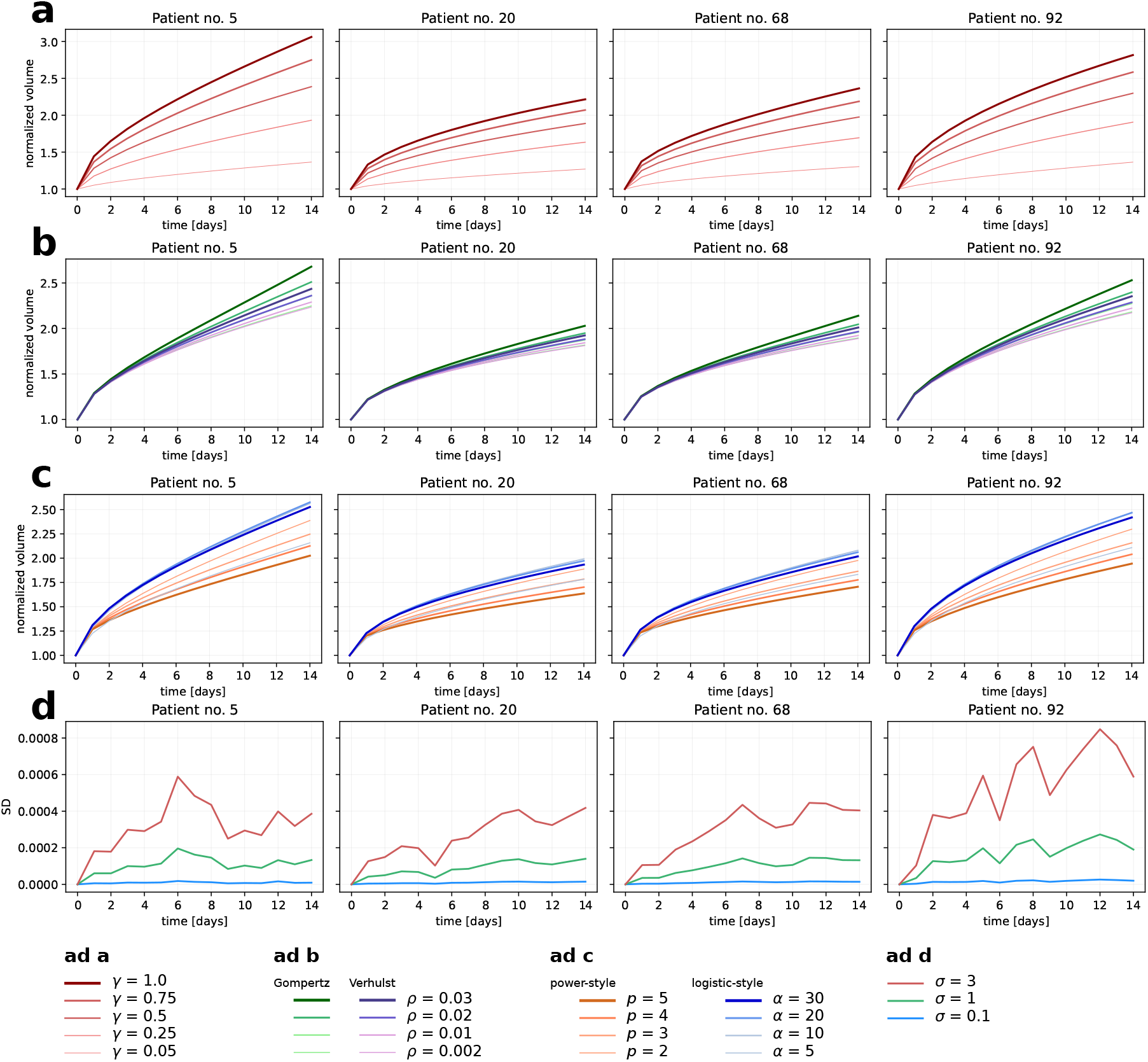
Parameter sensitivity analysis: varying *γ* (**a**), *ρ* and growth function (**b**), decay functions with their parameters *p* and *α* (**c**), and the stochastic parameter *σ* effect on volume SD (**d**). Volumes are normalized by their initial values. No additional scaling by FA. Abbreviations: FA (fractional anisotropy), SD (standard deviation).

Generally, the plots indicate that the value of *γ* is the most important in determining the dynamics of the GBM invasion, as it has the most significant impact on GBM tumor volume. Using the Gompertz growth function leads to a higher or equal volume in comparison to the Verhulst growth function. The decay functions show a more varied behavior, as the volume is consistently the highest when using the logistic-style decay function. The spread of trajectories is noticeably lower in the case of a logistic-style decay function, which means that the parameter *α* is generally less sensitive. On the other hand, the parameter *p* of the power-style decay function significantly affects the dynamics, with higher values of *p* leading to lower volumes. The SDs behave proportionally to *σ*, because the overall inter-trial variability across volumes grows faster and the individual steps are harder to predict as *σ* increases. In addition, some differences between patients can be observed. For example, the cases of patients no. 5 and 92 increase their relative volume more than the cases of patients no. 20 and 68.

### 4.4 Ablation experiment: low-grade glioma

Although the proposed model is formulated for GBM, its behavior can be further explored by attempting to model glioma types characterized by very different pathology dynamics, such as low-grade glioma (LGG). Quantitative MRI data analyses have recently shown that LGGs do not spread along white matter tracts (Rauch *et al*., 2025), while GBMs do (Esmaeili *et al*., 2018). Because the degree of white matter tract utilization can be directly modified via the *γ* parameter, the LGG behavior can be approximated by setting *γ* ≈ 0. Setting *γ* = 0 completely deactivates the spread (which would not be true for LGGs), as the graph weights obtained from connectome processing are the only transportation mechanism present in the model. Consequently, the LGG approximation via strongly suppressed *γ* represents an interesting ablation experiment, as direct integration of data from tractography constitutes the most novel aspect of the proposed model.

Side-by-side comparisons of the GBM and pseudo-LGG simulation results are provided in Figure 7. The 28-day pseudo-LGG simulation was repeated 5 times, other parameters were the same as in Section 4, with the exception of *γ* = 0.001 (i.e., 0.1% utilization of the connectome by the tumor). No additional FA scaling was performed. It can be seen that the GBM simulations are using white matter tracts to micro-infiltrate faster and farther into the surrounding tissue via many tentacle-like extensions, while the pseudo-LGG simulations do not exhibit this behavior and instead spread slowly locally with only subtle anisotropy. It must be reiterated that the exact underlying structural connectome used by GBM simulations is still present and used by the pseudo-LGG simulations, but in an almost completely suppressed manner.

**Figure 7:**
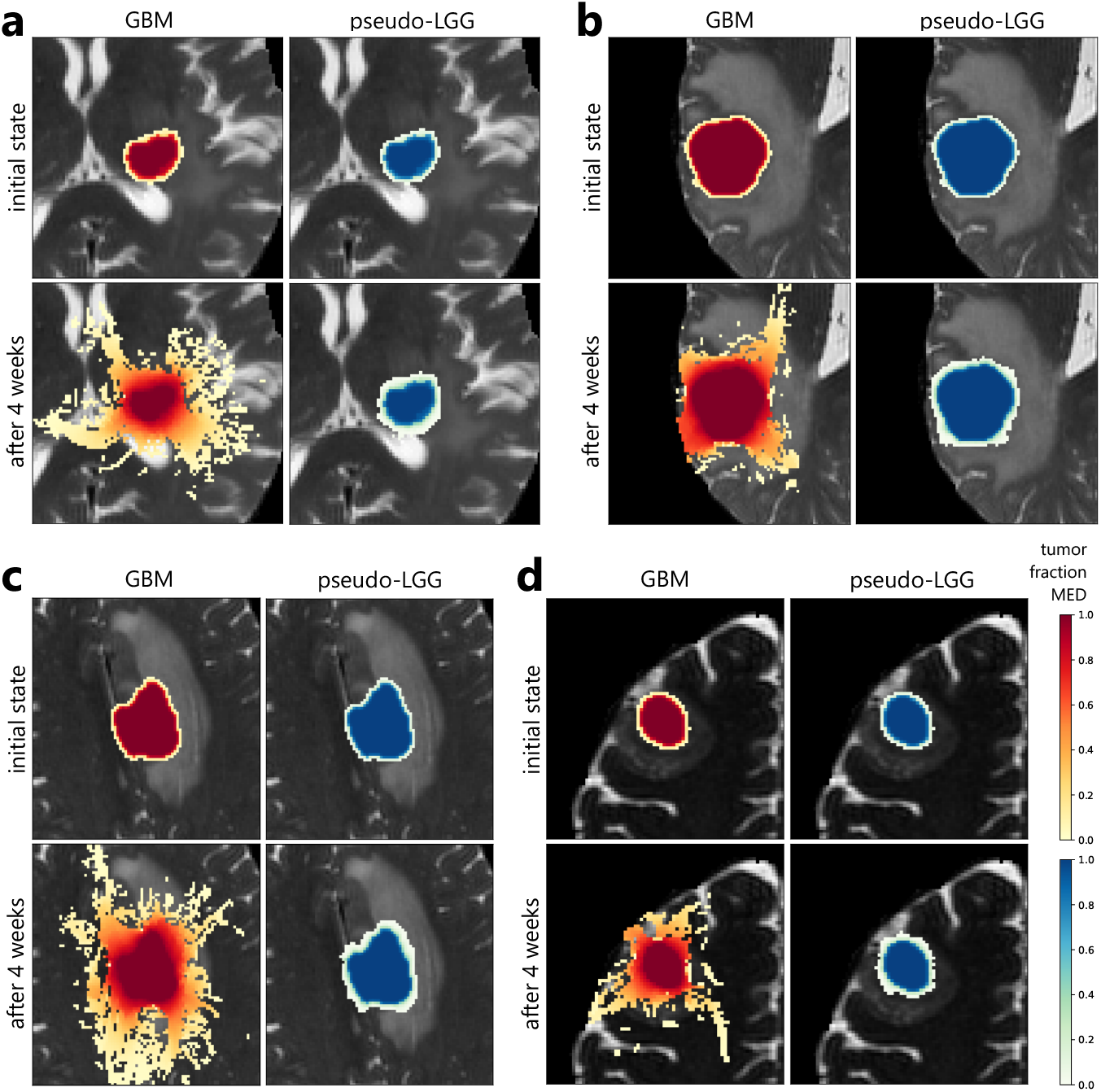
Comparison of GBM (yellow-to-red colors) and pseudo-LGG simulations (green-to-blue colors): patient no. 5 (**a**), 20 (**b**), 68 (**c**), and 92 (**d**). No additional FA scaling. Abbreviations: FA (fractional anisotropy), GBM (glioblastoma), LGG (low-grade glioma), MED (median).

## 5 Discussion

The GBM growth simulations reflect previously described invasion patterns. In the case of patient no. 5, the model correctly reflects previous observations that CST is highly susceptible to invasion by GBM and that it is structurally affected by it (Fekonja *et al*., 2021; Wang *et al*., 2024). In the case of patient no. 20, the simulated invasion of large white matter tracts is in accordance with observations by Wang *et al*. (2020). In the case of patient no. 68, the estimate of GBM invasion seems to indicate the rare butterfly GBM subtype, which describes tumors that spread to both hemispheres through the corpus callosum and are associated with a poorer prognosis and require different treatment strategies (Bjorland *et al*., 2022). In the case of patient no. 92, the invasion of SLF may be very likely, as spatial working memory deficits were previously observed in patients with right prefrontal glioma, which was linked to SLF damage (Kinoshita *et al*., 2016). Although the simulations reflect previously observed patterns of infiltration, the predictive ability of the model should be tested using longitudinal data in the future.

Using additional edge weight scaling by FA resulted in spatially distinct GBM invasion preferences. For example, the application of FA-based scaling exhibited a preference to spread along the commissural fibers in patient no. 5, while an increased growth of the central tumor mass was observed in patient no. 20. FA is used as a proof-of-concept example, but scaling the graph using other biomarkers derived from MRI and scaling methods presents an interesting area for future experiments.

The parameter sensitivity analysis shows that changing the degree of structural connectome utilization through *γ* has the most important role in GBM volume evolution. Changing the decay function resulted in a greater effect on the volume dynamics in comparison to changing the growth function. The stochasticity increases consistently with higher *σ* values.

The ablation experiment shows that the tentacle-like extensions emerge only if the white matter tracts are used by the tumor to micro-infiltrate, and otherwise either do not emerge at all, or after a significant time delay. Consequently, one of the main proposed model contributions, the graph-based non-linear diffusion spread encoded in the diffusion term (12), is the primary driving force behind the emergence of complex and far-reaching micro-infiltrative extensions from the GBM core.

Based on the provided examples, it can be stated that the model exhibits fast micro-invasion, while the central tumor mass grows more slowly. This behavior, although biologically grounded (Dada *et al*., 2026), is also likely a consequence of the initial tumor state, which only contains the observable tumor core. In reality, micro-infiltration is already advanced, but it is impossible to initialize due to the stated lack of sufficiently sensitive neuroimaging modality.

Due to physical factors or discretization imprecisions, some imperfections will always be present in the imaging data, such as partial-volume effect, registration distortions, interpolation and movement artifacts, etc. Consequently, the tractography algorithm can be misguided in tracing the structural brain connectome used as the 3D transportation network, especially when considering additional distortions caused by the tumor mass on the surrounding white matter structures. Nevertheless, this noise, although unwanted, might actually contribute to making the GBM growth less well-behaved, introducing implicit structural noise. The model may be extended by integrating probabilistic tractography to express the structural noise more explicitly.

The model simulates micro-infiltration on a macroscopic scale by treating the GBM tumor as a relatively homogeneous mass that is spreading, while the biological reality at the cellular level is much more complex and heterogeneous. Although similar simplifications are common in mathematical models, the reasoning behind adapting the macroscopic scale is grounded in real-world limitations and concerns. In order for the model to be applicable in clinical practice, it should first rely on *in vivo* imaging data that can be collected routinely without significant overhead for imaging departments. The degree of necrosis present is important for treatment planning, tumor classification, and patient prognosis, but the exact segmentation of various tissue types is rarely done aside from research purposes. Nevertheless, finding a way to connect both macroscopic and cellular-level approaches into one integrated model may lead to a more biologically plausible model.

The model is structurally parametrized by node and edge weights, but also by constants. The structural parameterization is performed by processing patient-specific MRI data, making the underlying transportation network strictly individual and personalized, but the constants are not tuned for each patient separately. Optimizing these constants to reflect patient-specific dynamics is one of the crucial next steps in development. Potential directions leading to a parameter optimization procedure consisting of ML, processing of multimodal MRI data, and inclusion of various genetic and pathological biomarkers associated with GBM progression.

Comparison with other existing models is challenging. They are rarely accompanied by the full source code and the mathematical descriptions are not always fully transparent, meaning that replicating the models requires a significant amount of trial-and-error testing. The MRI data used by the studies are also typically inaccessible, making it impossible to evaluate whether the replicated model behaves as it should. In addition to scientific transparency, the goal of using publicly available data and providing the full source code is to facilitate the inclusion of the proposed model in future benchmarking studies.

The proposed model can be used to estimate future GBM recurrences further away from the resection cavity, since GBM growth is explicitly modeled along white matter tracts, which were quantitatively shown to guide tumor cell invasion during recurrence (Shimizu *et al*., 2026). In addition, disruptions in the structural connectome caused by growth of the GBM can be studied to better link tumor progression with cognitive decline in patients with the GBM. Focusing on the GBM invasion morphology based on anatomical localization can also be an interesting follow-up study, as the exemplar simulations showed significantly varied infiltration patterns. The authors believe that the proposed transition from 3D voxel grids to graphs will facilitate further research of the relations between GBM dynamics and the brain connectome. Additionally, the model could be combined with ML approaches to perform various tasks, some of which are listed by Kukrál (2026).

Future studies should experiment with integrating other imaging modalities into the model. However, introducing other modalities should be guided by biomedical knowledge and not just to increase the complexity of the model for the sake of it. Although the model is intended for GBMs, it might be modified to describe other pathologies, as illustrated in Section 4.4. Similarly, isotropic growth could be achieved through weighting the graph’s edge weights by the same constant, new compartments could be introduced by adding more weights to the graph, etc. Interestingly, the model might be applicable with some changes to tumors outside of the brain as well, assuming that they exhibit strong directional spread that can be reliably reconstructed from the imaging data. However, such cases were not assessed and are purely hypothetical.

## 6 Conclusion

In this study, a novel mathematical model of GBM spread has been proposed and tested using MRI data from real clinical cases of patients with histopathologically confirmed GBM. The model expands on the well-established class of reaction-diffusion glioma growth models by formulating the diffusion mechanism on a weighted graph constructed from patient-specific diffusion MRI data and by introducing a stochasticity term corresponding to environmental noise imposed on GBM proliferation. The proposed model excels at modelling tentacle-like micro-infiltrative extensions from the tumor core, which was shown by the ablation experiment to be directly tied to the newly introduced diffusion along the tractography-weighted graph. Additional edge weight scaling using FA was tested, as well as the effect of individual model parameters on tumor volume dynamics. The deliberate choice was made to use the publicly available UCSF-PDGM dataset and open-source software tools to ensure maximal transparency and reproducibility in accordance with the principles of open science.

## Data Availability

The UCSF-PDGM dataset can be freely downloaded from the TCIA website. The complete source code and shareable derived data are available on GitHub. Links to both are:

https://www.cancerimagingarchive.net/collection/ucsf-pdgm/

https://github.com/kukrma/glioblastoma-connectome-invasion-model

## Code and data availability

The complete source code and shareable derived data are available on GitHub at https://github.com/kukrma/glioblastoma-connectome-invasion-model.

The UCSF-PDGM dataset can be freely downloaded from the TCIA website at https://www.cancerimagingarchive.net/collection/ucsf-pdgm/.

## Acknowledgments

The authors thank RNDr. Joná š Volek, Ph.D. and RNDr. Vladimír Š ví gler, Ph.D. from the Department of Mathematics (University of West Bohemia in Pilsen) for their valuable feedback on the mathematical formulation of the model, which helped clarify and improve its description.

This research received no specific grant from any funding agency in the public, commercial, or not-for-profit sectors.

## Author contributions

**Martin Kukrál:** Conceptualization, Data curation, Formal analysis, Investigation, Methodology, Software, Validation, Visualization, Writing – original draft, Writing – review & editing. **Roy A.M. Haast:** Writing – review & editing. **Irena Holečková:** Supervision, Writing – review & editing.

